# Circulating CD4+ T cells in people with HIV and history of pulmonary tuberculosis have more intact HIV DNA

**DOI:** 10.1101/2024.03.04.24303502

**Authors:** Marc Antoine Jean Juste, Yvetot Joseph, Dominique Lespinasse, Alexandra Apollon, Parmida Jamshidi, Myung Hee Lee, Maureen Ward, Esther Brill, Yanique Duffus, Uche Chukwukere, Ali Danesh, Winiffer Conce Alberto, Daniel W. Fitzgerald, Jean W. Pape, R. Brad Jones, Kathryn Dupnik

## Abstract

**Background:** The primary barrier to curing HIV infection is the pool of intact HIV proviruses integrated into host cell DNA throughout the bodies of people living with HIV (PLHIV), called the HIV reservoir. Reservoir size is impacted by the duration of HIV infection, delay in starting antiretroviral therapy (ART), and breakthrough viremia during ART. The leading infectious cause of death worldwide for PLHIV is TB, but we don’t know how TB impacts the HIV reservoir.

**Methods:** We designed a case-control study to compare HIV provirus-containing CD4 in PLHIV with vs. without a history of active TB disease. Study participants in the pilot and confirmatory cohort were enrolled at GHESKIO Centers in Port au Prince, Haiti. Intact and non-intact proviral DNA were quantified using droplet digital PCR of PBMC-derived CD4 cells. For a subset, Th1 and Th2 cytokines were assayed in plasma. Kruskal-Wallis tests were used to compare medians with tobit regression for censoring.

**Results:** In the pilot cohort, we found that PLHIV with history of active pulmonary TB (n=20) had higher intact provirus than PLHIV without history of active TB (n=47) (794 vs 117 copies per million CD4, respectively; p<0.0001). In the confirmatory cohort, the quantity of intact provirus was higher in the TB group (n=13) compared with the non-TB group (n=18) (median 102 vs. 0 intact provirus per million CD4, respectively p=0.03). Additionally, we found that the frequencies of CD4+ T cells with any detectable proviral fragment was directly proportional to the levels of IL1B (p= 0.0025) and IL2 (p=0.0002).

**Conclusions:** This is the first assessment of HIV provirus using IPDA in a clinical cohort from a resource limited setting, and the finding of larger reservoir in PLHIV with history of TB has significant implications for our understanding of TB-HIV coinfection and HIV cure efforts in TB-endemic settings.

## MANUSCRIPT

The primary barrier to curing HIV infection is the pool of intact HIV proviruses integrated into host cell DNA throughout the bodies of people living with HIV (PLHIV), called the HIV reservoir. Reservoir size is impacted by the duration of HIV infection, delay in starting antiretroviral therapy (ART), and breakthrough viremia during ART. Increased reservoir size has been associated with worse clinical outcomes, including neurocognitive impairment (1). The leading infectious cause of death worldwide for PLHIV is TB, but we don’t know how TB impacts the HIV reservoir. An approximation of reservoir size can be attained by quantitative PCR of HIV DNA in peripheral blood mononuclear cells (PBMC). One group used this approach to study HIV DNA levels in PBMCs from PLHIV with and without active pulmonary TB in Uganda and did not see a statistically significant difference (2). However, in a cohort in China, PLHIV with history of TB had a higher level of circulating HIV DNA than people without TB, but they also had higher viral load pre-ART (3). People with TB-HIV coinfection may have increased risk of virological failure and breakthrough viremias, resulting from or contributing to greater quantities of HIV DNA in circulating CD4+ T cells.

The emerging gold standard for the measurement of HIV reservoirs is the intact proviral DNA asay (IPDA), which distinguishes intact from defective proviruses, the latter being much more numerous (4). We applied a modified IPDA (5) to a cohort of people with HIV in a resource-limited setting with a TB syndemic to determine how quantities of CD4+ T cells containing HIV provirus vary in people who had had active bacteriologically confirmed pulmonary TB disease concurrent with or after diagnosis of HIV.

Using IPDA (*see supplemental methods)*, we measured proviral loads for 100 PLHIV participating in a TB study at GHESKIO Centers in Port au Prince, Haiti, 50 people with active or history of bacteriologically confirmed pulmonary TB and 50 people with no history of TB (Supplemental Table 1). Within the total cohort, there was a nearly 8-fold difference in intact provirus levels between current or history of TB vs. no TB (median 881 vs 116 copies per million CD4+ T cells, respectively; p=0.0001) (Figure 1A). Tobit regression of detectable intact provirus also showed statistically significant difference in intact provirus (p < 0.0001) between the TB and non-TB groups. There was no statistically significant difference in intact provirus between people with newly diagnosed untreated TB (n=25) vs. history of TB or TB currently being treated (n=25) (median 897 vs. 865 copies per million CD4+ T cells, respectively; p=0.92; Supplemental Figure 1A). In the people with no history of TB, the amount of intact provirus in CD4+ T cells was not statistically significantly different by IGRA status (median 80 for IGRA-positive vs.118 for IGRA-negative per million CD4, p=0.65; Supplemental Figure 1B).

**Figure 1.**
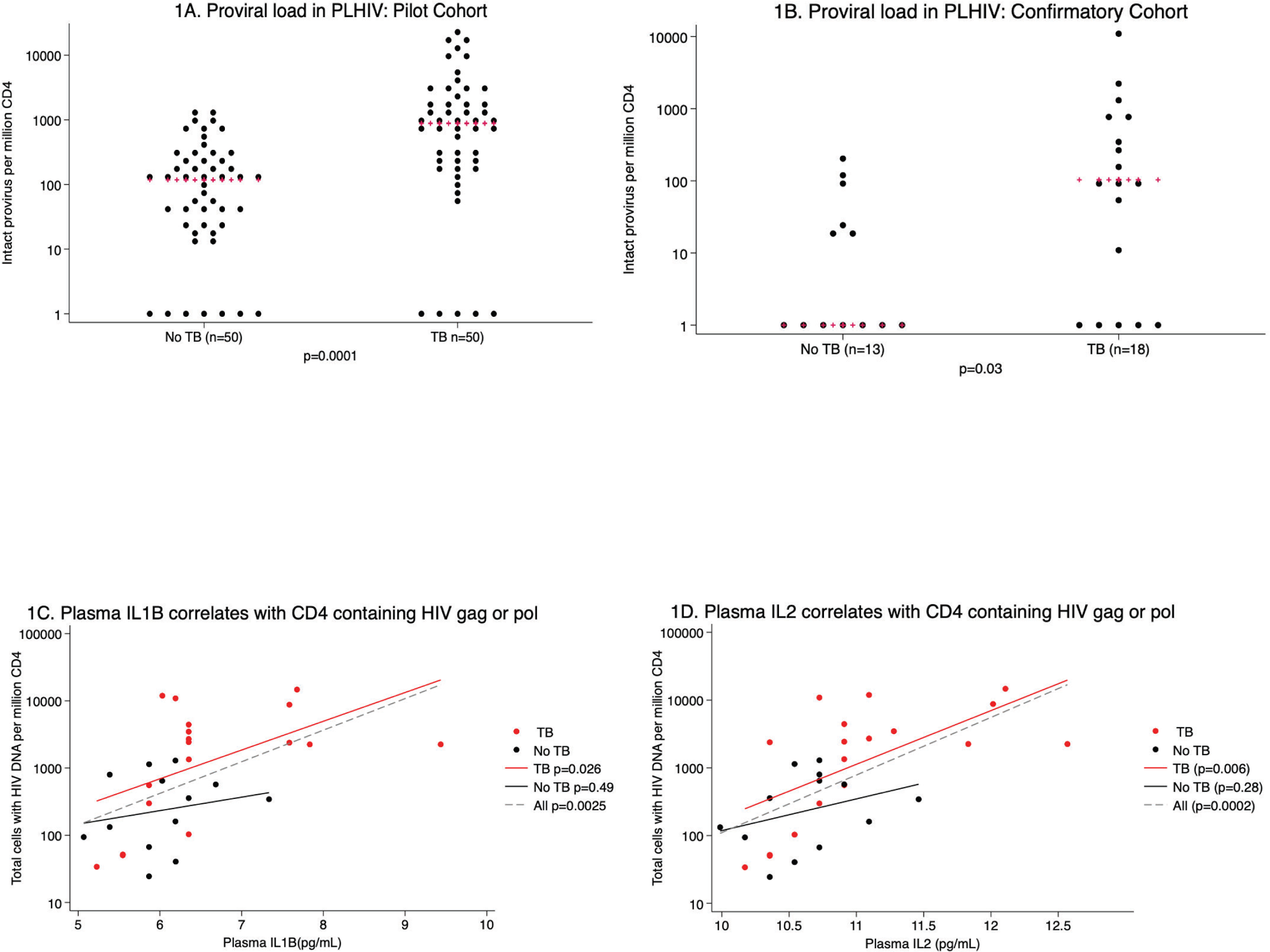
There was approximately 8-fold higher circulating CD4 T cells with intact HIV provirus in PLHIV with TB or TB history compared with no history of TB (p=0.0001, 1A). This was also found in a confirmatory cohort (p=0.03, 1B). In the confirmatory cohort, plasma IL1B (1C) and IL2 (1D) levels correlated with circulating CD4 T cells containing HIV gag and/or pol (p=0.0025 and p=0.0002, respectively).

To remove potential confounders of the difference in intact provirus observed in the initial groups, we excluded 26 people with HIV diagnosed less than one year before sample collection. We excluded 3 people with no plasma available for p24 testing and 3 people with detectable plasma p24. One person who had TB prior to HIV diagnosis was also excluded. After these adjustments, the difference in intact proviral load between TB and no-TB groups remained statistically significant (median 794 vs 117 copies per million CD4, respectively; p=0.0001) by Kruskal-Wallis test as well as by tobit regression (p<0.0001). This difference remained statistically significant when analyzing women and men independently (p< 0.005).

For the confirmatory study, we began with 34 participants from a study of TB recurrence. One participant’s PBMC did not have enough live cells after thawing to have CD4+ T cells selected and DNA extracted so was excluded from downstream analyses. One DNA sample failed QC with no detectable intact, *env* fragment, or *psi* fragment with all tested primer sets and so was excluded. One person had HIV viral load > 1,000 copies/mL as well as detectable p24 after dissociation. This person was excluded as the high-level viremia could have rendered the IPDA quantitation inaccurate. In the final analysis group from the confirmatory cohort (Supplemental Table 2), the quantity of intact provirus was higher in the TB group compared with the non-TB group (median 102 vs. 0 intact provirus per million CD4, respectively p=0.03) (Figure 1B). We also analyzed the intact provirus levels using a tobit regression model with 12 left-censored observations for intact proviral loads less than the lower limit of detection and the difference remained statistically significant (p=0.04). In the participants with no history of TB, there wasn’t a statistically significant difference in intact proviral load with IGRA positivity (median 0 vs 9 intact provirus per million CD4, p=0.96; Supplemental Figure 2A). In participants with history of one or recurrent episodes of TB, there was no statistically significant difference in intact proviral load in CD4+ T cells (median 103 vs 103 intact provirus per million CD4, p=0.55; Supplemental Figure 2B).

Additionally, we found that the frequencies of CD4+ T cells with any detectable proviral fragment was directly proportional to the levels of IL1B (0.0025, Figure 1C), IL2 (p=0.0002, Figure 1D), IL12p70 (p=0.0093), and IL13 (p=0.0067). After Bonferroni correction for the 8 analyzable cytokines, (cutoff p_adj_=0.05/8=0.006), the association between plasma IL-1B and IL2 and CD4 containing HIV *gag* or *pol* remained statistically significant. The difference in IL1B and IL2 levels between the TB and no-TB groups was not statistically significant (p=0.08 and p=0.1, respectively), but by observation of the fit lines for the cytokine levels by proviral load, the levels were overall higher for the TB group than for the non-TB group (Figure 1C and 1D).

In our pilot and confirmatory cohorts, we documented significantly higher levels of intact HIV proviruses in circulating CD4+ T cells of PLHIV with histories of bacteriologically confirmed pulmonary TB. Levels of HIV DNA correlated directly with levels of the pro-inflammatory cytokines IL1B and IL2 with a trend to higher levels in people with a history of TB. Strengths of the study include the use of IPDA to discriminate intact versus defective HIV proviruses, translational approach using samples from people who have experienced the true natural history of HIV-TB coinfection, and confirmatory cohort testing. Limitations include incomplete CD4+ T cell count nadir data because of the changing landscape of CD4 testing in HIV care, inclusion of only bacteriologically confirmed pulmonary TB, and HIV proviral load testing at only a single time point. This is the first assessment of HIV provirus using IPDA in a clinical cohort from a resource limited setting, and the finding of 8-fold larger reservoir in PLHIV with history of TB has significant implications for our understanding of TB-HIV coinfection and HIV cure efforts in TB-endemic settings.

PLHIV with TB exhibit systemic inflammation and immune activation. Local inflammation has the propensity to increase HIV replication and viral load at sites where HIV and replicating Mtb exist, even prior to the development of symptomatic TB (6). Mtb cell wall components are able to activate TLR signaling to induce HIV replication without live bacteria and may induce HIV transcription in neighboring cells that are uninfected with Mtb (6). These factors may contribute to the larger HIV reservoir in PLHIV with history of pulmonary TB, and should make TB a serious consideration for all reservoir-targeting HIV cure endeavors.

## Data Availability

All data produced in the present study are available upon reasonable request to the authors after publication.

## 1. Supplemental materials and methods

### Sex as a biological variable

This study included men and women. A post-hoc analysis with stratification by sex was performed which showed similar findings of statistically significantly higher percentage of CD4+ T cells containing intact provirus in people with history of pulmonary TB.

### Study site

The prevalence of HIV in adults in Haiti was 1.8% in 2021. Haiti has the highest rate of TB in the Americas with 154 new cases of TB per 100,000 people in 2022, of whom 15% were HIV-positive (7). GHESKIO centers in Port au Prince, Haiti is a Haitian non-profit organization that is the largest provider of HIV and TB care in the Americas.

### Clinical cohorts

For the pilot study, people with microbiologically confirmed pulmonary TB were enrolled from TB ambulatory care centers at GHESKIO as part of a study on tuberculosis. People living with HIV with or without history of TB were enrolled from the TB and HIV clinics at GHESKIO. Latent TB testing was using the Quantiferon-TB Gold In-Tube interferon-gamma release assay (IGRA) (Qiagen, MD, USA). Blood for peripheral blood mononuclear cell (PBMC) isolation was collected with heparin as an anticoagulant. The pilot nested case-control study included people with new diagnosis of bacteriologically confirmed pulmonary TB, people with history of bacteriologically confirmed pulmonary TB, and people with no history of TB who were either IGRA positive or IGRA negative.

For the confirmatory study, volunteer participants were enrolled in a study of the human immune response to TB with an emphasis on recurrent TB at GHESKIO. Participants had HIV and were receiving suppressive ART with a history of zero, one, or two episodes of TB after HIV diagnosis. They had one study visit with questionnaire, demographics, and phlebotomy. Latent TB testing was using the Quantiferon-TB Gold Plus In-Tube interferon-gamma release assay (IGRA) (Qiagen, MD, USA). Blood for peripheral blood mononuclear cell (PBMC) isolation was collected with EDTA as an anticoagulant.

For both studies, PBMC were isolated at GHESKIO using density gradient separation (Ficoll-Hypaque, GE Healthcare) with 5×10^6^ cell aliquots frozen and then shipped to New York where they were stored at −135C. PBMC specimens were identified with a unique patient identifier which did not correspond to their TB status. For analysis, participants were assigned to TB and no-TB groups after laboratory assessments were complete.

### Intact proviral DNA assay (IPDA)

For each participant, one aliquot of 5×10^6^ cells was thawed and washed before CD4+ T cell isolation by negative selection (EasySep^TM^ Human CD4+ T cell Enrichment Kit, Stem Cell Technologies). DNA was then isolated using a column-based method (DNeasy Miniprep Kit, Qiagen) and quantitated using Nanodrop (Thermo Fisher Scientific). Intact proviral DNA amplification was determined using droplet digital PCR (ddPCR) on a QX200 instrument (Bio-Rad) (4). Amplification of the HIV *gag* (*psi*) and *pol* regions and human *rpp30* were done independently in parallel with quadruple technical replicates and for DNA shearing using *rpp30*. PCR reactions with fewer than 10,000 droplets read were excluded from the calculation for that participant. Participants who did not have any detectable *env* or *psi* amplification using the primers and conditions described in Bruner et al (4) and Gunst et al (8) were re-run with secondary *env* primers and probes as described in Kinloch et al (5). Participants with no amplifiable HIV DNA (*gag* fragment and *psi* fragment) with both primer sets were also excluded (n=1). From the ddPCR, we generated numbers of 3’ defective (*env*), 5’ defective (*psi*), and presumed intact provirus containing both *env* and *psi* per million CD4+ T cells.

### HIV viral load

Blood from participants in the pilot TBRU study were collected with heparin as the anticoagulant, so viral load results were not reliable. Instead, we tested plasma for HIV circulating p24 gag protein using an ELISA (RETRO-TEK™ HIV-1 p24 Antigen ELISA, ZeptoMetrix, Buffalo, NY) with immune dissociation and reactive confirmation (HIV-1 p24 ICx/CRx kit, ZeptoMetrix). Participants who had detectable p24 or who did not have plasma available for p24 testing were excluded from final analysis of the pilot cohort.

Participants in the TB recurrence study were to have had undetectable viral loads prior to enrollment in the study. However, because of the impact that active HIV replication could have on the IPDA, we measured p24 in these participants as well using plasma collected with EDTA anticoagulant. Plasma was centrifuged at 10,000 xg for 10 minutes to remove particulates. RNA was extracted from plasma using a semi-automated method and HIV viral load quantitated using a previously described integrase single copy assay (9, 10). Participants who had viral load > 1,000 copies/mL and detectable p24 after immune dissociation and neutralization were excluded. Samples were analyzed on an ABI Viia7 Real-Time PCR System. Cycle threshold values were compared with a validated HIV RNA standard run to determine concentration.

### Cytokine levels

Plasma levels of granulocyte-macrophage colony-stimulating factor, interferon gamma, interleukin 1 beta (IL1B), interleukin 2 (IL2), interleukin 4 (IL4), interleukin 5 (IL5), interleukin 6 (IL6), interleukin 12p70 (IL12p70), interleukin 13 (IL13), interleukin 18 (IL18), and tumor necrosis factor (TNF) were measured using a bead-based multiplex assay (Th1/Th2 11-plex Human ProcartaPlex Panel, Invitrogen). Quantitation was performed using a Luminex MAGPIX system with xPONENT v4.3 software (Luminex, Austin, TX) according to the manufacturer’s instructions with the following specifications. Plasma was spun at 10,000 xg for 10 minutes to remove particulates prior to aliquoting onto the plate. Incubation was overnight at room temperature. All samples were run in duplicate and the average reading used for quantitation with the standard curve. For samples that were less than the lower limit of detection, we imputed a value of half the difference between zero and the lowest concentration standard on the standard curve for each cytokine. Cytokines that were less than the level of detection for more than 50% of plasma samples did not continue to statistical analyses.

### Data and Statistical analysis

Demographic and clinical characteristics were expressed as numbers and percentages with interquartile range given for continuous variables. All statistical analysis was done in Stata (version 18, College Station, TX, USA). Distributions of variables were tested using Skewness/Kurtosis tests for normality. For normality test p< 0.05, Kruskal Wallis tests were used to compare medians and for normality test p> 0.05, two-sided t-tests were used for comparison of means. For proviral loads reported by IPDA as being less than the lower detection level, we input a level of 1 copy per million CD4 for two reasons: 1) the fact that these participants have uncured HIV means that their level can’t be zero and 2) we analyzed proviral loads using log_10_ because of the large numerical distribution of proviral loads and log_10_0 is undefined whereas log_10_1 is zero. When there were a large number of undetectable (log_10_1=0) proviral loads, we utilized tobit regression for left censoring of proviral loads that were less than the lower limit of detection with the IPDA (11). After completing the Luminex plasma cytokine assay, we excluded cytokines for which more than half of participants had a level less than the lower limit of detection. We applied Bonferroni correction for multiple analyses using the remaining number of cytokines as the correction factor. We used Pearson’s correlation coefficient (<pwcorr> in Stata) to compare quantities of provirus and cytokines. For all analyses, the cutoff for significance was p≤0.05.

For a post-hoc analysis, we compared the rates of usage of alternative primers in the IPDA to determine if differences in proviral loads between TB and non-TB groups were related to use of different primer sets. The rates of primer usage by group are included in Supplemental Tables 1, 2, and 3. For the first cohort, where there was a statistically significant difference in use of alternate primers between the groups, we stratified by primer type and compared log intact proviral load between TB groups separately in those samples amplified by original IPDA primers (p=0.0012) and those samples amplified by alternate primers (p=0.016).

### Study approval

This study was approved by institutional review boards at Weill Cornell Medicine and GHESKIO Centers. Study participants provided written informed consent. All participants were offered antiretroviral and anti-TB treatment and continuing care for HIV and TB per the Haitian national guidelines and the GHESKIO standard of care.

### Data availability

The data used to generate the figures and supplemental figures are included in the Supporting Data Values document. Additional information is available from the authors upon reasonable request.

## 2. Financial Contributions

The authors recognize funding to KD (NIH K23-AI131913 and R01-AI176943, Doris Duke Charitable Foundation Clinical Scientist Development Award) and DWF (NIH U19-AI111143, P30-AI168433, and K24-AI098627). Additionally, this project utilized databases maintained in REDCap, supported by UL1-TR002385 from the National Center for Advancing Translational Sciences of the NIH. We acknowledge the patient care, clinical research, and laboratory teams at GHESKIO Centers in Port au Prince, Haiti.

## 3. Supplemental references

7. WHO. Tuberculosis profile: Haiti.

8. Gunst JD, Pahus MH, Rosas-Umbert M, Lu IN, Benfield T, Nielsen H, et al. Early intervention with 3BNC117 and romidepsin at antiretroviral treatment initiation in people with HIV-1: a phase 1b/2a, randomized trial. Nat Med. 2022;28(11):2424-35.

9. Cillo AR, Vagratian D, Bedison MA, Anderson EM, Kearney MF, Fyne E, et al. Improved single-copy assays for quantification of persistent HIV-1 viremia in patients on suppressive antiretroviral therapy. J Clin Microbiol. 2014;52(11):3944-51.

10. McCann CD, van Dorp CH, Danesh A, Ward AR, Dilling TR, Mota TM, et al. A participant-derived xenograft model of HIV enables long-term evaluation of autologous immunotherapies. J Exp Med. 2021;218(7).

11. Tobin J. Estimation of Relationships for Limited Dependent-Variables. Econometrica. 1958;26(1):24-36.

## 4. Author Contributions

MAJJ*– clinical studies (TBRU – pilot cohort), acquire data

YJ* – clinical studies (TB recurrence – confirmatory cohort), acquire data

DL – clinical studies (TB recurrence – confirmatory cohort), acquire data

AA – technical expertise

PJ – conduct experiments, write manuscript

MHL – analyze data

MW – conduct experiments, acquire data

EB– conduct experiments, acquire data

YD– conduct experiments, acquire data

UC – conduct experiments, acquire data

AD– conduct experiments, acquire data

WCA– conduct experiments, acquire data

DWF – design research studies, data interpretation

JWP – design research studies

RBJ– design research studies, analyze data, data interpretation

KMD – design research studies, conduct experiments, acquire data, analyze data, data interpretation, write manuscript

---

**Supplemental Table 1.** Demographics of the pilot cohort.

|  | TB group<br>(n=50) |  | No TB group<br>(n=50) |  |
| --- | --- | --- | --- | --- |
|  | Untreated TB<br>(n=25) | History of TB or on treatment<br>(n=25) | IGRA-positive<br>(n=25) | IGRA-negative<br>(n=25) |
| Sex (#female (%)) | 10 (40%) | 14 (56%) | 16 (64%) | 17 (68%) |
| Age in years (mean +/- SD) | 38.16 +/- 7.5 | 38.8 +/- 6.8 | 44 +/- 5.9 | 44 +/- 4.3 |
| Time since HIV dx in years<br>(median (IQR)) | 0.02 (0.02-0.03) | 3.1 (1.1-5.4) | 12.8 (11.7-14.3) | 11 (10.5-14.7) |
| Time on ART in years (median<br>(IQR)) | 0 | 2.9 (0.9-4.5) | 10 (8.7-10.7) | 8.9 (8-10.2) |
| Time between diagnosis and<br>ART (median (IQR)) | .06 (0.02-0.07) | 0.9 (0.4-.57) | 2.7 (2.3-4.1) | 2.6 (1.3-4.7) |
| Alternate IPDA primers used<br>(number (%)) | 7 (28%) | 9 (36%) | 4 (16%) | 3 (12%) |

**Supplemental Table 2.** Demographics of the final analyzed group in the confirmatory cohort.

|  | TB group<br>(n=18) | No TB group<br>(n=13) |
| --- | --- | --- |
| Sex (#female (%)) | 10 (55%) | 7 (54%) |
| Age (mean +/- SD) | 44 +/- 10.1 | 43.3 +/- 12.1 |
| Time since HIV dx in years (median (IQR)) | 6.7 (5.4, 7.8) | 6.4 (4.7, 7.9) |
| Time on ART in years (median (IQR)) | 6.2 (4.1, 7) | 5 (4, 6.5) |
| Time between diagnosis and ART (median (IQR)) | 0.1 (0, 0.2) | 0.8 (0, 1.6) |
| HIV viral load (median, (IQR)) | 0 (0-518) | 0 (0-77) |
| Alternate IPDA primers used (number (%)) | 4 (22.2%) | 3 (23%) |

**Supplemental Figure 1.**
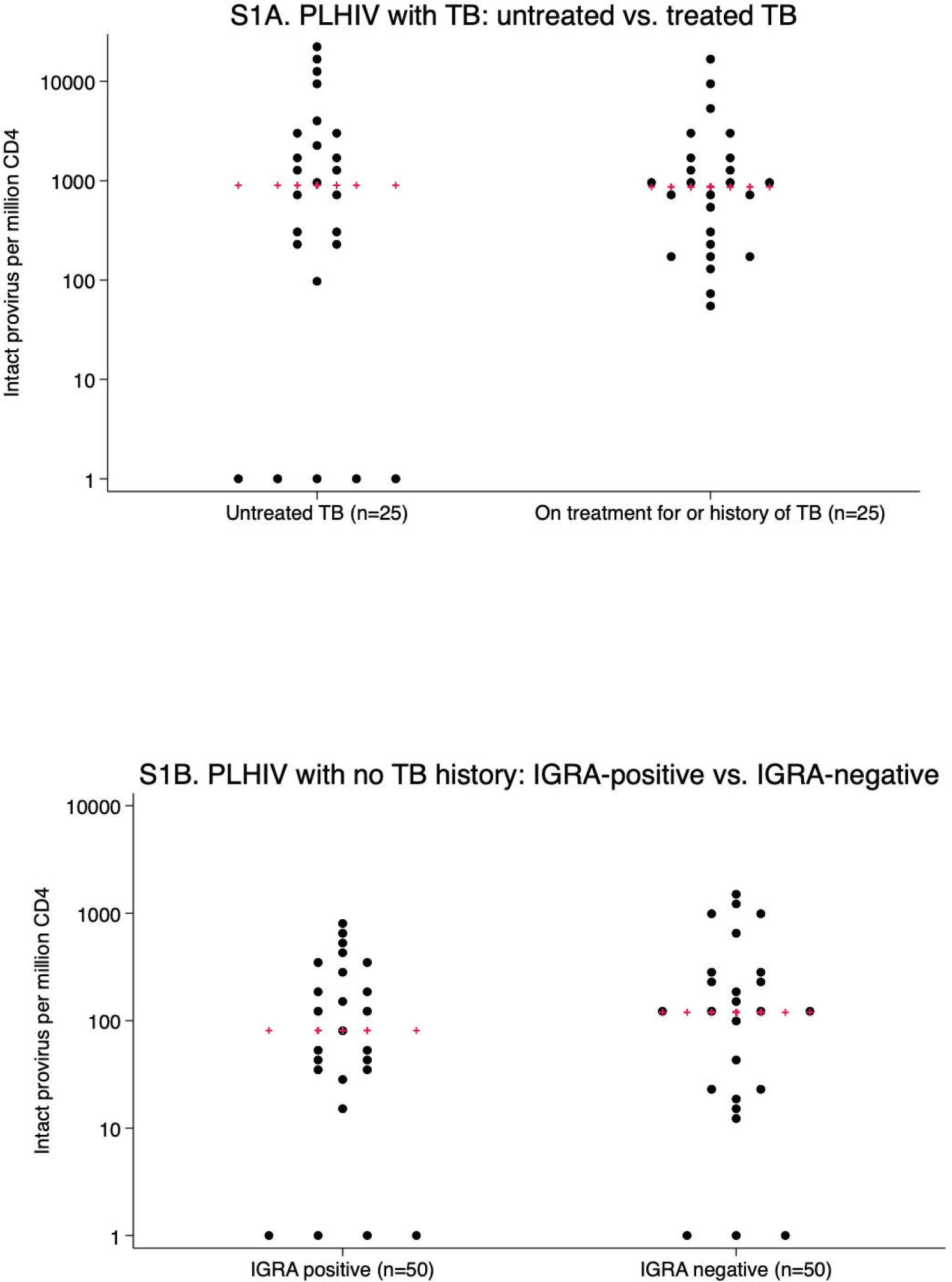
Subgroup analysis in the full pilot cohort of intact proviral load in circulating CD4 T cells. There was no statistically significant difference between intact provirus levels for people with active vs treated pulmonary TB disease (p=0.92, S1A) or by IGRA positivity in people with no history of active pulmonary TB (p=0.96, S1B). Red pluses are the medians.

**Supplemental Figure 2.**
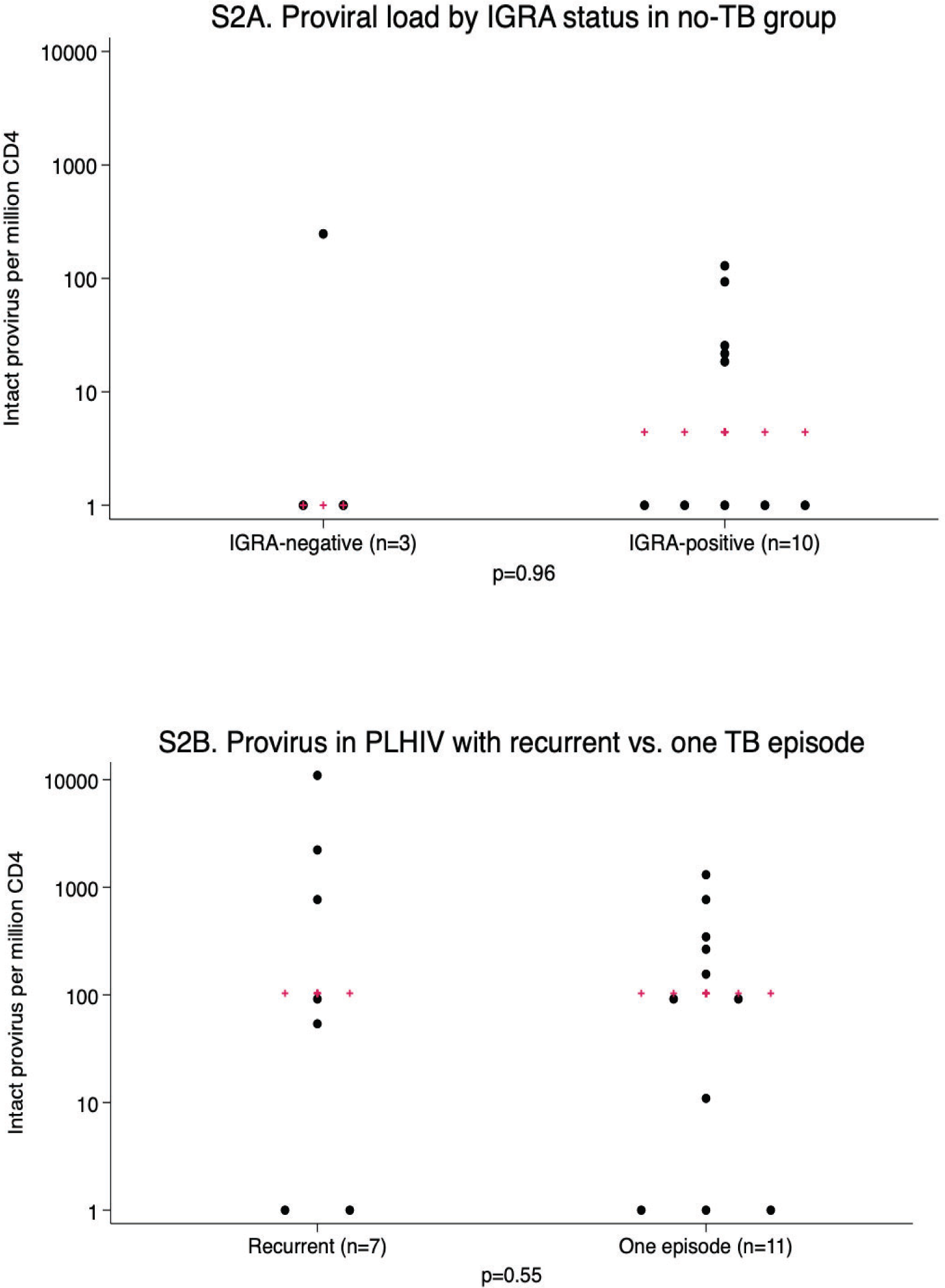
There was no statistically significant difference in intact provirus in people without TB with different IGRA status at the time of study enrollment (S2A). There was also no statistically significant difference in intact provirus in people with TB history with recurrent vs only one episode of TB (S2B). Red pluses are the medians.

